# Effort aversion and diminished exploration in apathy associated with Traumatic Brain Injury

**DOI:** 10.1101/2025.08.22.25334260

**Authors:** Jeremy Hogeveen, Ethan Campbell, Denicia Aragon, Ebony Pearson, Caitlin Enders, John Romero, Lauren Brown, Richard A. Campbell, Darbi Gill, Davin K. Quinn, Masud Husain, Andrew R. Mayer, Vincent D. Costa

## Abstract

Humans often prefer familiar, safe rewards to avoid the cognitive effort required to explore novel options. Here, we examined patients with motivational deficits following traumatic brain injury (TBI) as a clinical model to understand the neural mechanisms driving this trade-off. Participants with TBI and healthy controls performed *two* model-based fMRI tasks: i) a physical effort-value tradeoff task, and ii) an explore-exploit decision-making task. While diminished motivation (i.e., apathy) in both groups was associated with a devaluation of prospective reward based on physical effort, apathy in TBI patients was associated with a distinct deficit in the motivation to explore novel opportunities. Specifically, while TBI patients with acquired apathy sometimes selected novel options, they failed to adequately weight the latent future value of the information that these options provided. This indicates that they explored less effectively, as their choices were less guided by the informational value of novel options. This inability to track the “exploration bonus” of novel options in acquired apathy was also linked to impaired reward prediction error (RPE) encoding in key decision-making circuitry, including prefrontal cortex and the striatum. These findings suggest that RPE serves as a critical teaching signal, helping to shape the motivational drive to engage in directed exploration.

## Background

Traumatic brain injury (TBI) can lead to chronic and disruptive new-onset psychiatric symptoms^1^. These often include a reduced motivation to engage in goal-directed behaviors—commonly known as apathy^2–4^. Apathy in TBI has been decoupled from traditional assays of injury severity and time since injury^3^, and instead appears to be driven by complex disruptions to distributed brain networks that are moderated by compounding psychosocial factors, ultimately shaping the trajectory of recovery and leading to lasting motivational deficits in a subset of patients^5^. While clinically recognized as a major impediment to recovery and quality of life^4,6^, the precise cognitive and neural mechanisms of apathy remain under debate^5^. An unresolved question is whether apathy stems from an aversion to effort, a general impairment in value-based learning, or a deficit in the drive to explore novel opportunities for future benefit.

Recent years have seen considerable advances in understanding the neural mechanisms of motivated decision-making in normative samples^7–9^. Yet, there is a comparative lack of basic neuroscience research into the mechanisms that elevate risk for amotivation following a TBI event. Applying insights from decision neuroscience to clinical populations is critical for identifying meaningful dissociations that can explain complex symptoms like apathy^2,10–12^. For example, a decreased willingness to exert physical effort for rewards is a well-documented feature of apathy, observed in both patients with Parkinson’s Disease (PD) and in subclinical apathy in healthy individuals (CTRLs)^13,14^. There also may be some basic decreases in reward sensitivity and reward-based learning associated with clinical apathy in both PD^15,16^ and TBI^2,17,18^. Notably, exploiting choices with high, well-learned reward values requires relatively low cognitive effort. In contrast, tracking uncertainty in the reward environment and deciding when it is appropriate to seek out novel information—a form of ‘directed exploration’^19,20^—requires its own form of effort. This distinction is critical, as a reduced motivation to explore novel options that may lead to greater future rewards has not been studied in TBI-related apathy. Aberrant explore-exploit decision-making is an emerging target for understanding both normative and pathological motivational states, with deficits in clinical populations providing meaningful insights into reward processing and prefrontal-subcortical circuit disruptions^21–26^. Therefore, it remains unclear if apathy in TBI is driven primarily by an aversion to physical effort, impaired reward-based learning, or a specific impairment in the motivation to engage in directed exploration of prospective rewards.

There are several lines of evidence suggesting that clinical apathy may affect the exploration of new opportunities. For example, when healthy participants are given dopamine agonists prior to performing a ‘Patch Foraging’ task they more readily explore new locations when the expected value in their current location is low^27^. This finding directly mirrors the impact of dopamine agonists on novelty-directed exploration in nonhuman primates (NHPs)^28^. Relatedly, patients with PD-related apathy demonstrate more ‘random’ exploration and impaired value learning on a task where learning to exploit familiar choice options is cognitively demanding^16^. Degeneration or pharmacological manipulation of dopaminergic circuitry may therefore modulate motivated behavior not only by shifting effort-based decision-making for rewards, but also by changing one’s motivation to gain new information about the environment through exploration.

To investigate these possibilities, we used functional MRI (fMRI) and two distinct decision-making tasks in healthy volunteers and patients with chronic TBI. To probe physical effort-based decision-making, we used an ‘Apples Task,’ where participants decided whether to exert varying levels of physical effort for different reward magnitudes^14^. To examine the trade-off between exploiting familiar rewards based on their known value versus exploring new possibilities to reduce uncertainty, we used a ‘Novelty-Bandit Task’^24,25,29^. The Novelty-Bandit required choices between familiar options with a known reward history and novel options with uncertain future value. Because prior studies have rarely examined exploration alongside the exploitation of well-learned values, they may have missed a key functional dissociation relevant to understanding clinical apathy. We hypothesized that—in line with prior studies in PD and CTRLs—reduced willingness to exert physical effort for rewards would represent a general, transdiagnostic dimension of apathy. We also hypothesized specific dimensions of apathy in TBI would be associated with deficits in cognitively effortful directed exploration, which may or may not be related to subclinical apathy in CTRLs.

Our findings support these hypotheses. While a greater reluctance to expend physical effort for rewards was linked to apathy across TBI patients and CTRLs, a reduced motivation to explore was specifically associated with apathy in the TBI group. This was tied to a blunted reward prediction error signal in key motivational brain regions in TBI patients with increased apathy—suggesting that this neurocomputational signal is linked to the motivation to engage in directed exploration. These results provide a more precise framework for understanding acquired amotivation, pointing to potential new avenues for treating a symptom that profoundly interferes with daily life in TBI.

## Methods and Materials

### Participants

A total of *N*=70 participants (*N*=49 chronic TBI patients, and *N*=21 non-brain-injured healthy controls (CTRLs)) were initially recruited. Of those enrolled, *N*=17 (*N*=15 TBI and *N*=2 CTRL) participated during an initial pilot phase prior to collection of the Apathy Motivation Index (AMI). Given the AMI was the central apathy measure in the current study, these early participants were excluded due to the enrollment timeline, rather than symptom severity, data quality issues, or other unexpected dropout. This yielded a total sample of *N*=53 participants (*N*=34 TBI, *N*=19 CTRL) with complete clinical and AMI data (characterized in **Tables 1-2**). Prior to task analysis and fMRI modeling, *N*=5 additional participants (*N*=4 TBI, *N*=1 CTRL) were excluded due to having one or more data quality issues—insufficient responding during the decision-making tasks (*N*=3), absence of confirmed loss of consciousness during the structured TBI interview (*N*=2; see **Semi-structured TBI interview** section), and/or excessive head motion during one or both fMRI tasks (*N*=2)^1^. This resulted in a final task and fMRI analysis sample of *N*=48 participants (*N*=30 TBI patients and *N*=18 CTRLs). Importantly, within the main TBI cohort, the *N*=4 excluded TBI participants did not differ from the final sample included in the primary behavioral and fMRI analyses with respect to apathy (AES: *U*=60.5, *p*=1.0), post-concussive symptoms (NSI: *U*=58.5, *p*=0.98), age (*U*=78.5, *p*=0.36), or TBI severity (0.25 versus 0.27 msTBI; Fisher’s exact *p*=1.0). Because the TBI and CTRL groups differed in age (**Table 1**), age was included as a covariate in second-level fMRI models, and examined as a control variable in additional post-hoc behavioral models. The UNM Health Science Center’s Human Research Protections Office approved the study and all participants provided informed consent.

**Table 1.** Final sample characteristics. TBI and CTRL columns report mean +/- SD except where otherwise indicated. Inferential tests comprised Welch’s *t*-tests for comparisons that were normally distributed but with unequal variance between groups, and Mann-Whitney *U*-tests for comparisons that were not normally distributed. Alternative hypothesis for each test is that CTRL<TBI. Acronyms: NSI (neurobehavioral symptom index), BDI (Beck depression inventory), BAI (Beck anxiety inventory), BIS (Barratt impulsivity scale), and FD (framewise displacement).

|  | TBI | CTRL | INFERENTIAL TEST |
| --- | --- | --- | --- |
| *N* | 34 | 19 | N/A |
| **Age** | **39.7+/-10.7** | **28.1+/-10.08** | ***U*=143.5, *p*<0.001** |
| *Female (prop.)* | 0.42 | 0.38 | $\chi^2=0.076, p=0.782$ |
| *Years of Ed. (mean+/-SD)* | 15.3+/-2.84 | 16.2+/-2.76 | *U*=341.5, *p*=0.212 |
| **NSI** | **27.2+/-19.2** | **5.83+/-5.93** | ***U*=74.0, *p*<0.001** |
| **BDI** | **13.2+/-10.43** | **4.5+/-3.97** | ***U*=127.5, *p*<0.001** |
| **BAI** | **17.1+/-11.76** | **6.5+/-8.15** | ***U*=107.0, *p*<0.001** |
| **BIS** | **67.4+/-11.6** | **55.6+/-7.73** | ***t*=4.32, *p*<0.001** |
| *Bandit FD* | 0.24+/-0.16 | 0.18+/-0.06 | *U*=284.0, *p*=0.239 |
| *Effort FD* | 0.25+/-0.14 | 0.21+/-0.07 | *U*=294.0, *p*=0.427 |

**Table 2.** Apathy measurements. Descriptive and inferential stats from the apathy evaluation scale (AES) and apathy motivation index (AMI) separated by group. Note: *Med*=median; *95%-CI*=95%-high density interval; *P*+=Proportion of posterior estimates above 0. *: *P+*≥0.95 or ≤0.05; **: *P+*≥0.99 or ≤0.01; ***: *P+*≥0.999 or ≤0.001; *NS*: *P+*<0.95 or >0.05. Effect sizes were computed with CTRL as the reference (i.e., median *d* values are negative when apathy is higher in TBI).

| AES Behavioral | Med=9.1<br>CI=8.3-10 | Med=8.0<br>CI=7.0-9.2 | $d=-0.49$<br>$CI=-1.2$ to $0.17$; $P+=0.07$ |
| --- | --- | --- | --- |
| AES Cognitive* | Med=14<br>CI=12-15 | Med=12<br>CI=10-13 | $d=-0.54$<br>$CI=-1.1$ to $0.05$; $P+=0.04$ |
| AES Emotional | Med=3.9<br>CI=3.4-4.4 | Med=4.0<br>CI=3.4-4.6 | $d=0.06$<br>$CI=-0.53$ to $0.64$; $P+=0.59$ |
| AES Other | Med=5.7<br>CI=5.0-6.5 | Med=5.5<br>CI=4.6-6.4 | $d=-0.13$<br>$CI=-0.74$ to $0.48$; $P+=0.33$ |
| AES Total (Mean) | Med=8.2<br>CI=7.4-9.0 | Med=7.4<br>CI=6.6-8.2 | $d=-0.40$<br>$CI=-0.99$ to $0.18$; $P+=0.09$ |
| AMI Behavioral* | Med=1.8<br>CI=1.5-2.0 | Med=1.3<br>CI=0.91-1.7 | $d=-0.69$<br>$CI=-1.4$ to $0.007$; $P+=0.02$ |
| AMI Emotional* | Med=1.3<br>CI=1.1-1.5 | Med=1.0<br>CI=0.79-1.3 | $d=-0.49$<br>$CI=-1.1$ to $0.11$; $P+=0.05$ |
| AMI Social | Med=1.7<br>CI=1.4-1.9 | Med=1.4<br>CI=1.1-1.7 | $d=-0.41$<br>$CI=-1$ to $0.18$; $P+=0.09$ |
| AMI Total (Mean)* | Med=1.6<br>CI=1.4-1.7 | Med=1.3<br>CI=1.1-1.5 | $d=-0.72$<br>$CI=-1.4$ to $-0.09$; $P+=0.01$ |

### Bayesian Modeling

Bayesian modeling approaches and posterior distribution-based inferences were used throughout the current manuscript. To ensure clarity, posterior distributions were summarized using the median, and uncertainty was quantified using 95% credible intervals (*95%-CIs*). The direction of effects was evaluated using *P+*, representing the posterior probability that a parameter is strictly positive. We interpreted *P+* ≥ 0.90, ≥ 0.95 as moderate or strong evidence for a positive effect, and *P+* ≤ 0.10 or ≤ 0.05 as moderate or strong evidence for a negative effect, respectively.

### Clinical Assessments

#### TBI assessment

##### Pre-screening

Participants were pre-screened by phone to confirm they did (TBI) or did not (CTRL) have a history of brain injury. All TBI subjects were in the late chronic post-injury phase at enrollment (≥6 months). The median time since injury across the sample was 110.47 months (*SD*=83.41 months).

##### Semi-structured interview

Participants completed a modified version of the Rivermead interview to determine TBI history^30,31^. Participants included in the CTRL sample reported no TBI history. In the TBI group, only participants reporting ≥1 TBI event involving a loss of consciousness (LOC) were included in the final sample. Several patients reported more than one TBI (*N*=20/34). The majority of the TBIs were classified as mild [mTBI; LOC<30 minutes^32^; *N*=26/34]. A subset of patients reported at least one moderate-to-severe TBI (msTBI; LOC>30 minutes^32^; *N*=8/34). Repeated versus single mTBI was not associated with incidental findings on structural MRI (*Χ*^2^=0.05, *p*=0.82), while msTBI patients *were* more likely to have a structural MRI finding (*Χ*^2^=6.98, *p*=0.008).

##### Clinical assessments

We administered the Neurobehavioral Symptom Inventory (NSI^33^) to assess the type and severity of post-concussive symptoms, as well as the Beck Depression Inventory (BDI^34^), Beck Anxiety Inventory (BAI^35^), and Barratt Impulsiveness Scale (BIS) to provide additional measures of psychiatric symptoms post-injury. We measured both the Apathy Evaluation Scale (AES^36^) and the Apathy Motivation Index (AMI^37^) to assess dimensions of amotivation in the current sample. AES was included for consistency with the prior literature, but AMI was considered the primary apathy outcome as it has been rigorously psychometrically evaluated. The AMI provides measures for three subdomains of apathy (behavioral activation, emotional sensitivity, and social motivation), which can also be combined to generate a total apathy score.

### Decision-Making Tasks

Stimulus presentation and timing during fMRI scans were controlled using EPrime Version 3 (Psychology Software Tools, Sharpsburg, PA), and manual responses were recorded using the MIND Input Device (https://www.mrn.org/collaborate/mind-input-device).

#### Apples task

We used a physical effort-based decision-making for reward task, modeled after one that has been used extensively in previous studies of behavioral apathy in healthy participants and neurological patients^13,14^ (**Figure 1A**). Prior to fMRI, participants made a series of 10s duration grips of a dynamometer at varying levels of maximum effort (i.e., percentage maximum voluntary contraction, % MVC). Levels were either 20, 40, 60, 80, or 100% MVC, and each grip intensity was practiced for 10s with a roughly 20-30s inter-grip rest interval. Participants were then told that during the fMRI task they were going to make a series of effort-value tradeoff decisions based on the levels of % MVC that they had just practiced. On each choice trial, effort levels were 20, 40, 60, 80, or 100% MVC, and reward value ranged from 1-13 points (5 levels: 1, 4, 7, 10, and 13). Each Effort*Value combination was repeated three times per fMRI run (75 trials per run), repeated across two runs (150 total trials).

**Figure 1.**
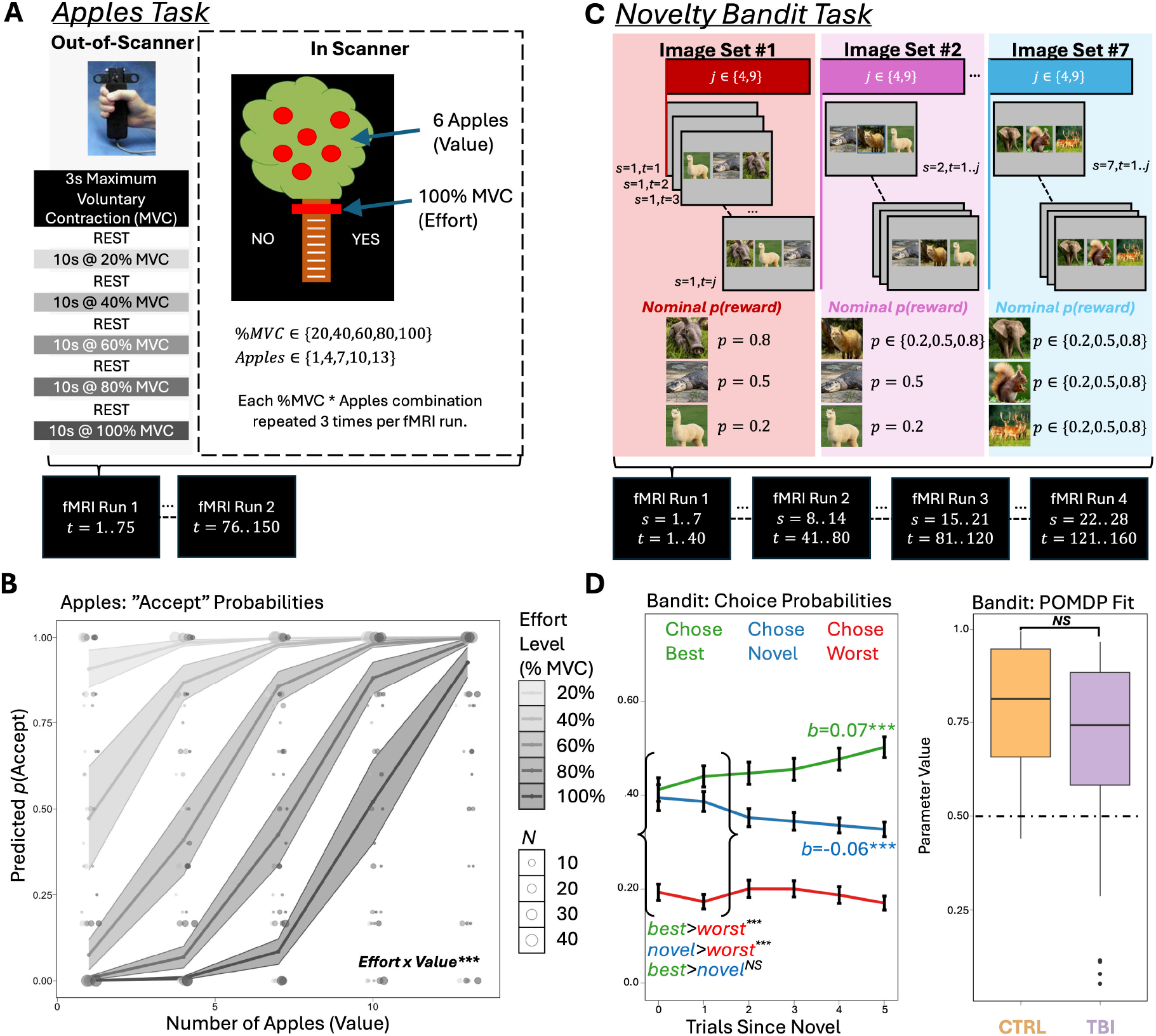
Overview of fMRI Tasks and Derived Measures. **(A)** Apples Task: An effort-based decision-making task that trained participants to exert grips at varying percentages of their maximum voluntary contraction (MVC), then required them to say whether they Accept or Reject various physical effort-reward value tradeoffs during fMRI (two 75-trial runs; 150 total trials). **(B)** There was a significant interaction between effort and value in predicting the probability of Accepting offers (*P+*=0.001), suggesting participants demonstrated an effort-based discounting effect—i.e., requiring higher reward values to Accept offers at higher levels of physical effort. **(C)** Novelty-Bandit Task: A probabilistic reinforcement learning task in which participants choose among three options with low, medium, or high reward probabilities. After several trials, a novel stimulus replaces one familiar option, a process that occurs seven times per fMRI run (four 40-trial runs; 160 total trials). **(D)** After novel insertions, participants were equally likely to explore (choose the novel option; *med*=0.39) or exploit (choose the best familiar option; *med*=0.42; NS: *P+*=0.84), and over time became increasingly exploitative (*b*=0.07, *P+*=1.0) and less exploratory (*b*=−0.06, *P+*=0.003). A normative partially observable Markov decision process (POMDP) model fit behavior well, with large effect size correlations between the model and observed choice behavior. This fit CTRL participants’ choices moderately better than TBI patients’ (*P+*=0.94). Notes: *: *P+*≥0.95 or ≤0.05; **: *P+*≥0.99 or ≤0.01; ***: *P+*≥0.999 or ≤0.001; *NS*: *P+*<0.95 or >0.05.

On each trial, participants had 3000 ms to respond ‘yes’ (i.e., Accept) or ‘no’ (i.e., Reject the effort-value tradeoff). Rather than have participants execute real dynamometer grips for each offer they accepted, responses were recorded with discrete button presses, to avoid the potential confound of summed BOLD activity driven by 10s isometric muscle contractions which would necessitate a lengthy return-to-baseline after each trial. Participants were instructed that they would be required to execute a random subset of the % MVC grips that they accepted after completion of the fMRI runs. There should be no reason to over- or under-estimate one’s willingness-to-exert physical effort in this setup, similar to the approach used extensively in studies of willingness-to-pay in value-based choice^38^. This approach recreated the canonical effort-based discounting effect on this paradigm (**Figure 1B**). Intertrial intervals were jittered (1000-7000ms) to optimize HRF deconvolution at the time of response.

#### Bandit task

##### Task description

We used a version of our Novelty-Bandit Task^25^ (**Figure 1C**). The Novelty-Bandit is a 3-armed probabilistic reinforcement learning task involving periodic novel stimulus insertions that cue participants to make explore-exploit decisions. On each trial (160 total trials), participants selected one of three neutral images from the International Affective Picture System (IAPS^39^). Participants completed four bandit task fMRI runs, each comprising 40 trials. At the start of each run, three images were selected at random and assigned a nominal probability of reward—low (*p*=0.2), medium (*p*=0.5), or high (*p*=0.8). Participants learned to select the most rewarding options based on positive (green ‘+1’) or neutral (black ‘−0’) feedback after each choice. The response window was fixed at 3000ms. After each response, there was a jittered inter-stimulus (ISI; 1000-4000ms) interval, and then feedback was presented for 1500 ms. Finally, a fixation cross was presented centrally for a jittered inter-trial interval (ITI; 1000-12000ms). ISI and ITI jitters were optimized separately enabling us to deconvolve the HRF at choice *and* feedback.

After ≈4-9 trials (randomized; *Median*=6) a novel stimulus was inserted in place of one of the familiar images, and assigned its own low, medium, or high reward probability. This occurred seven times per block, for 28 novel stimulus insertion trials across all fMRI runs. Participants were told they could earn an additional reward if they accumulated enough points in the task, ensuring a high level of motivation. Choice behavior in this task was similar to previous studies using the paradigm^22,25^ (**Figure 1D**).

##### Computational modeling

Optimal strategies on the Novelty-Bandit Task were fit using a partially observable Markov decision process (POMDP) model that has been described elsewhere^24,25^. Briefly, the POMDP estimates the value of all three options based on the summed likelihood that a particular action will be rewarded based on its reinforcement history (i.e., its Immediate Expected Value, IEV), and a latent estimation of the discounted sum of future rewards that can be obtained from the current set (i.e., the Future Expected Value, FEV). In the POMDP framework, the decision to exploit familiar rewards is motivated by maximizing IEV, whereas the decision to explore novel and uncertain choice options is motivated by an information bonus—i.e., a higher FEV for a given option relative to its alternatives (i.e., BONUS). This BONUS parameter is high for novel stimuli, but decreases over time once the option has been sampled. The POMDP model fit choice behavior well in both TBI (*median r*=0.69) and CTRL (*median r*=0.79), with a slightly higher correlation in CTRLs (*Median Difference*=0.10, *95%-CI*=−0.03 to 0.23, *P+*=0.94; **Figure 1D**).

### Magnetic Resonance Imaging Methods

#### Image acquisition and fMRIPrep preprocessing

Imaging data was acquired on a Siemens Prisma 3T system with a 32-channel phased-array head coil. T1-weighted structural images were acquired via multi-echo MPRAGE sequence (5-echo; voxel size=1mm iso). TBI participants also completed T2-weighted, FLAIR, and susceptibility-weighted imaging which were included for neuroradiology but not analyzed. Echo-planar imaging data was collected during the task fMRI runs with a TR=1s, TE=30ms, flip angle=44, multiband factor=4, and a voxel size of 3mm iso. Results included in this manuscript come from preprocessing performed using fMRIPrep 20.2.0rc0^40,41^ (RRID:SCR_016216), which is based on Nipype 1.5.1^42,43^ (RRID:SCR_002502). This fMRIPrep anatomical and functional preprocessing boilerplate for this version of the software is documented in full in Hogeveen et al., 2021^44^.

#### fMRI modeling

First- and second-level GLMs of the fMRI data were fit in FSL ^45^, and all first-level models included the standard 6 motion parameters and their derivatives (MP24^46^). On the Apples task, we modeled ‘Accept’ and ‘Reject’ events separately. These events were parametrically modulated by the ‘Value’ and ‘Effort’ of each offer. Our primary analysis from this task focused on the extent to which responses scaled as a main function of Effort, as well as the interaction between decision (Accept/Reject) and parametric modulators (Value/Effort). This allowed us to isolate brain signals that correspond to effort estimation (main effect of Effort), those corresponding to effort-discounted subjective value (e.g. scaling with value on ‘Accept’ trials and effort on ‘Reject’ trials), and those related to choice difficulty or conflict (e.g. scaling with effort on ‘Accept’ trials and value on ‘Reject’ trials). For the Bandit task, we used a model-based GLM with regressors derived from our POMDP. At the time of choice, key regressors included the value of exploiting (IEV) and exploring (BONUS). At the time of feedback, we modeled the reward prediction error (RPE), a key teaching signal known to influence explore-exploit decision-making^47^. Group-level analyses were conducted using FLAME1 with cluster-extent thresholding (*z*>2.3, *p*<0.05).

## Results

### Chronic Psychiatric Symptoms

Chronic TBI patients reported significant post-concussive symptoms, measured via the NSI (*p*<0.001; **Table 1**). Additionally, TBI patients reported higher levels of depression, anxiety, and impulsivity (all *p*s<0.001; **Table 1**). Repeated TBIs were associated with increased neurobehavioral symptoms (NSI—Repeated TBI: *M*=33.2±12.4; Single TBI: *M*=17.2±13.5; *t*=2.72, *p*=0.011), anxiety (BAI—Repeated TBI: *M*=21.3±12.4; Single TBI: *M*=10.1±6.29; *t*=3.39, *p*=0.002), and impulsiveness (BIS—Repeated TBI: *M*=70.7±10.3; Single TBI: *M*=61.8±11.7; *t*=2.20, *p*=0.039), but did not impact depression (BDI—Repeated TBI: *M*=15.1±11.2; Single TBI: *M*=10.0±8.41; *p*=0.155) relative to a single TBI. Psychiatric symptoms did not differ between individuals with mTBI vs. msTBI (all *p*s>0.259). This is consistent with evidence that repeated injuries can be more predictive of chronic post-concussive symptoms than TBI severity^48,49^. Apathy was observed across the spectrum of TBI in the current study^cf.,3,4,44^. On the AES apathy in TBI was higher on most dimensions relative to CTRL (**Table 2**; **Figure 2**). A similar pattern was observed on the AMI, with the largest effect sizes on the behavioral apathy subscale as well as the AMI total (**Table 2**; **Figure 2**). Within TBI, apathy scores were not impacted by repeated injury or severity.

**Figure 2.**
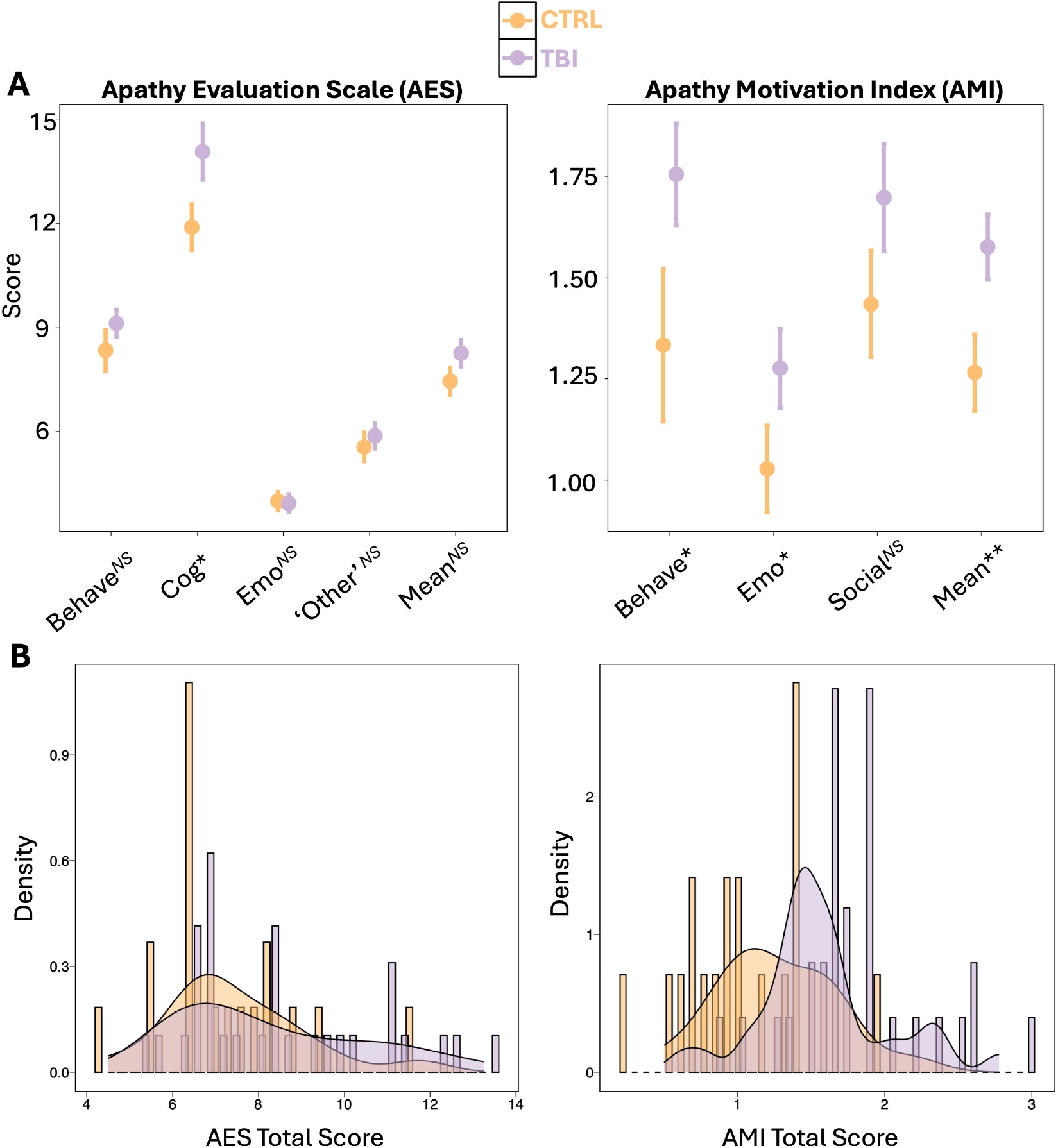
Apathy scales by group. **(A)** AES and AMI mean and standard error. TBI increased on AES cognitive, AMI behavioral, AMI emotional, and AMI total. **(B)** Distribution of apathy scores, averaged across AES and AMI subscales. Notes: *: *P+*≥0.95 or ≤0.05; **: *P+*≥0.99 or ≤0.01; ***: *P+*≥0.999 or ≤0.001; *NS*: *P+*<0.95 or >0.05.

### Effort-Based Discounting of Rewards

Bayesian multilevel logistic regression models were fit to responses (Accept=1; Reject=0) on the Apples task. In a main effects model, responses were fit as a function of apples (reward value), effort level, (%MVC), and group (TBI=1, CTRL=0), and a random intercept for subject. In an interaction model, we added a three-way interaction to determine whether the impact of effort on reward-based decision-making differs by group. There was statistical evidence for main effects of both effort (*b*=−0.96, *95%-CI*=-1.02 to −0.91) and reward value (*b*=0.33, *95%-CI*=0.32 to 0.35), and an interaction between reward and effort (*b*=0.03, *95%-CI*= 0.02 to 0.05; **Figure 1B**). There was no main effect of group—TBI and CTRL were similarly likely to accept offers (*b*=−0.29, *95%-CI*=-1.16 to 0.59). There was a three-way interaction between effort, value, and group (*b*=0.06, *95%-CI*=0.03 to 0.10). This interaction was driven by TBI patients discounting rewards more steeply at lower effort levels relative to CTRL.

We then determined whether apathy was associated with effort-based discounting of rewards in TBI and CTRL. To quantify individual differences in effort-based discounting, we fit a generalized linear mixed-effects model with a binomial link function to the trial-level response data from the Apples task with subject-specific random intercepts and slopes for the interaction between reward value and physical effort. We extracted random intercepts and slopes for the reward by effort interaction, to operationalize baseline acceptance probability across these parameters (intercept) and effort-based discounting of rewards (slope). Visual examination of model results revealed that individuals with a positive slope were willing to Accept the significant majority of offers when reward value was high, but individuals with a negative random slope remained unwilling to Accept at high effort levels irrespective of the available reward (**Supplemental Figure 1**). Therefore, more negative slopes reflected a greater degree of effort-based discounting of reward magnitude.

Both baseline acceptance (intercept) and effort discounting (slope) parameters were modeled as a function of total apathy, group, and their interaction. There was no statistical evidence that apathy, group, or their interaction affected baseline acceptance—indicating that these variables did not uniformly decrease or increase choice across levels of value and effort. There was a main effect of apathy on effort-based discounting (*b*=−0.18, *95%-CI*=−0.30 to −0.07). The main effect of group and the interaction term did not impact effort discounting. The main effects model was driven by a similar negative association, whereby increased apathy was associated with steeper effort-based discounting of rewards across *both* TBI (*ρ*=−0.43, *95%-CI*=−0.71 to −0.08, *P+*=0.01) and CTRL (*ρ*=−0.35, *95%-CI*=−0.72 to 0.11, *P+*=0.08) groups (**Figure 3A**).

**Figure 3.**
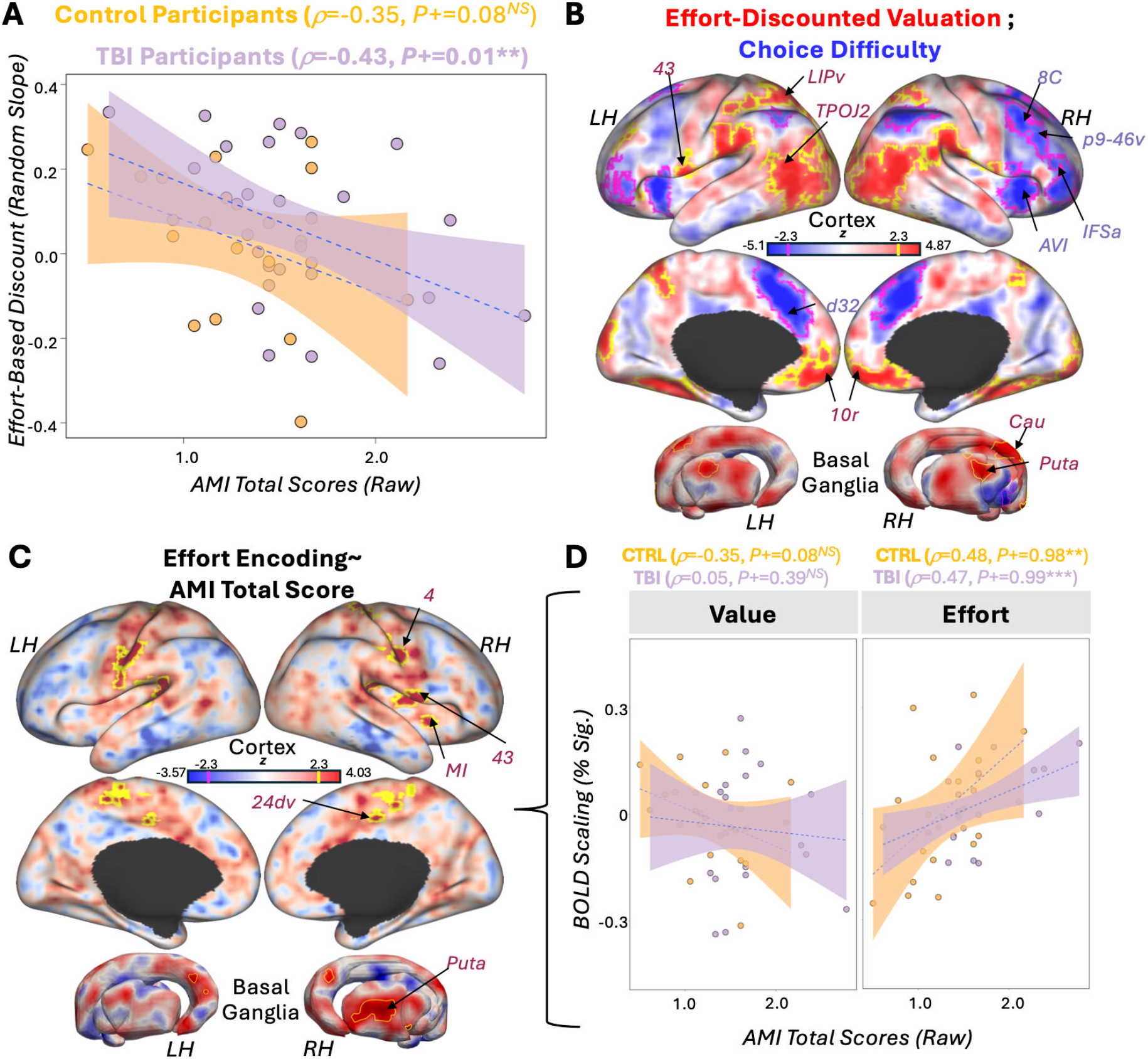
Effort-Based Discounting Correlates of Apathy Across TBI and CTRL. **(A)** Bayesian multiple regression models yielded credible evidence for a main effect of apathy on effort-based discounting of rewards—such that higher apathy scores are associated with greater degree of effort-based discounting. **(B**) Wholebrain map of the interaction between choice event (Accept versus Reject decisions) and relevant choice parameter (Value versus Effort). This revealed evidence for positive responses reflecting effort-discounted value encoding in ventromedial prefrontal cortex, temporo-parieto-occipital junction regions, lateral intraparietal area, and the dorsal striatum. Negative responses to this interaction term likely reflected the degree of choice conflict or difficulty and this was encoded in a large section of dorsal anterior cingulate cortex and dorsomedial prefrontal cortex, dorsolateral and ventrolateral prefrontal regions, and insular cortex. **(C)** Apathy total scores were positively associated with effort encoding across TBI and CTRL participants in a network including central somatomotor areas, midcingulate, middle insula, and putamen. **(D)** This association was driven by a credible positive correlation between apathy scores and BOLD scaling with the effort parameter across both groups. Notes: *: *P+*≥0.95 or ≤0.05; **: *P+*≥0.99 or ≤0.01; ***: *P+*≥0.999 or ≤0.001; *NS*: *P+*<0.95 or >0.05. ROI labels from the HCP multimodal parcellation (Glasser et al., 2016, *Nature*).

A wholebrain map revealed that effort-discounted value was encoded in ventromedial prefrontal cortex (vmPFC), lateral intraparietal cortex (LIP), temporo-parieto-occipital junction (TPOJ), subcentral area 43, and the putamen. Notably, in some regions (e.g. LIP, TPOJ, and putamen; **Figure 3B**) this was driven by a combination of increased Value encoding on Accept relative to Reject trials, and increased Effort encoding on Reject relative to Accept trials (**Supplemental Figure 2A**), whereas in others it was driven by either a Value encoding effect on Accept trials (e.g. vmPFC; **Supplemental Figure 2B**) or an Effort encoding effect on Reject trials (e.g. area 43; **Supplemental Figure 2C**). In contrast, choice difficulty was encoded in an extended anterior cingulate / dorsomedial frontal cluster, anterior and middle insula, and both dorsal and ventral regions of lateral prefrontal cortex (**Figure 3B**). In most of these regions this was driven by a combination of increased Value encoding on Reject trials and Effort encoding on Accept trials (**Supplemental Figure 2D**).

Across both TBI patients and controls, higher apathy was associated with increased BOLD signal that tracked the required effort level in a network of regions including pre- and post-central sulcal regions, subcentral area 43, middle insular area, and the putamen (**Figure 3C**). This effect was specific to effort encoding, not associated with value or the interaction between effort and choice. There was a robust positive correlation between AMI total scores and BOLD scaling with the effort parameter across choice types in both TBI (*ρ*=0.47,*P+*=0.99; **Figure 3D**) and CTRL (*ρ*=0.48,*P+*=0.98; **Figure 3D**) groups. Conversely, there was no strong relationship between apathy and BOLD scaling with the value parameter in either group in these ROIs, though CTRLs showed moderate evidence for a negative association (TBI: *ρ*=0.05, *P+*=0.39; CTRL: *ρ*=−0.35, *P+*=0.08; **Figure 3D**). This suggests that individuals with higher apathy exhibit a heightened neural sensitivity to effort, which may drive their behavioral tendency to discount prospective rewards more steeply as a function of physical effort costs.

### Explore-Exploit Decision-Making

After a novel stimulus was presented, participants were more likely to exploit the best available familiar option (*Med*=0.42, *95%-CI*=0.38−0.47) or explore the novel option (*Med*=0.39, *95%-CI*=0.34−0.44) than they were to select the worst familiar alternative (*Med*=0.18, *95%-CI*=0.15-0.21; Chose best versus chose worst: *d*=1.9, *95%-CI*=1.3 to 2.4, *P+*=1.0; Novel versus worst: *d*=1.6, *95%-CI*=1.1 to 2.2, *P+*=1.0; **Figure 1D**). Participants demonstrated a similar probability of exploiting (choosing the best alternative) and exploring (choosing the novel stimulus) on early choice trials (*d*=0.21, *95%-CI*=−0.20 to 0.63, *P+*=0.84; **Figure 1D**).

Beyond choice probabilities, we fit normative POMDP models of explore-exploit decision-making. The POMDP correlated well with behavior across TBI (*Med r*=0.69, *95%-CI*=0.59 to 0.79, *P+*=1.0; **Figure 1D**) and CTRL (*Med r*=0.79, *95%-CI*=0.70 to 0.87, *P+*=1.0; **Figure 1D**). Subject-level estimates for weighting the value of exploiting (IEV) and exploring (BONUS) options were derived via the POMDP, and used to test the hypothesis that explore-exploit behavior is driven by specific components of apathy. No main effects of apathy (on any dimension) or TBI were found for IEV or BONUS. However, two interactions emerged when predicting the BONUS parameter: both behavioral and emotional (but not social) apathy interacted with TBI group (**Figure 4A**). Within the TBI group, these interactions were driven by individuals with higher behavioral or emotional apathy demonstrating reduced motivation to explore when BONUS was high (AMI Behave: *ρ*=−0.36, *95%-CI*=−0.68 to −0.02, *P+=*0.03; AMI Emotion: *ρ*=−0.46, *95%-CI*=−0.73 to −0.11, *P+*=0.009), whereas residualized social apathy showed moderate evidence for an association in the opposite, positive direction (AMI Social: *ρ*=0.28, *95%-CI*=−0.10 to 0.61, *P+*=0.91; **Figure 4A**). BONUS was not associated with any AMI in CTRLs (behavioral AMI: *ρ*=0.05, *95%-CI*=−0.43 to 0.52, *P+*=0.58; emotional AMI: *ρ*=0.28, *95%-CI*=−0.18 to 0.70, *P+*=0.87; social AMI: *ρ* =0.16, *95%-CI*=−0.30 to 0.59, *P+*=0.74). There was moderate-to-strong statistical evidence that the behavioral and emotional apathy associations with exploration were stronger in TBI than in CTRLs (posterior difference, apathy-bonus correlation in CTRL versus TBI: behavioral AMI, *P+*=0.91; emotional AMI, *P+*=0.99). Notably, the associations between apathy domains and exploration bonus within the TBI group were not confounded by any extraneous variance associated with age, number of TBIs, TBI severity, or time since most recent TBI. Bayesian partial correlations demonstrated robust associations between AMI scores and bonus (AMI behavior ∼bonus: *ρ*=−0.45, *95%-CI*=−0.72 to −0.081, *P+*=0.012; AMI emotional ∼ bonus: ρ=−0.50, *95%-CI*=−0.77 to −0.19, *P+*=0.003) after regressing out the effect of these variables.

**Figure 4.**
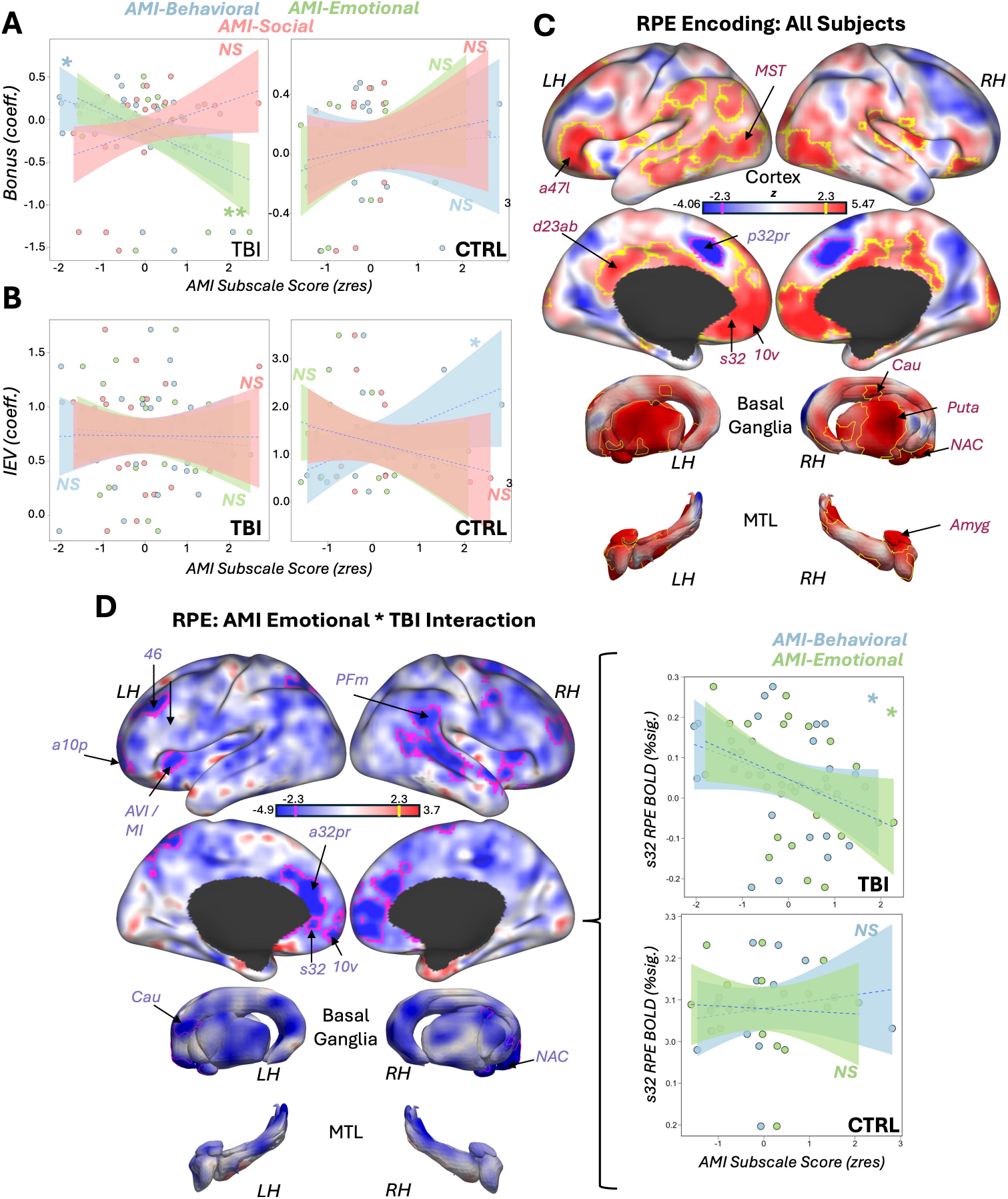
Novelty-Bandit Correlates of Acquired Apathy in TBI. **(A-B)** Bayesian multiple regression models yielded credible evidence for apathy by TBI group interaction on bonus, driven by negative associations between behavioral and emotional apathy and bonus in TBI, and no associations between apathy dimensions and bonus in CTRLs. There was a credible interaction between behavioral apathy and TBI group on IEV, with more apathetic CTRL participants demonstrating greater weighting of IEV, but no associations between apathy and IEV in TBI. **(C)** Wholebrain map of reward prediction error (RPE) encoding across both TBI and CTRL participants. Positive RPE encoding observed in ventromedial and ventrolateral prefrontal cortex, insula, ventral and dorsal stream visual areas, posterior cingulate, amygdala, and striatum. A region in dorsal anterior cingulate cortex was also found to negatively encode RPE. **(D)** There was a significant interaction between emotional apathy scores and TBI status on RPE BOLD encoding, driven by blunted RPE encoding in several areas of prefrontal cortex, insula, rostral anterior cingulate cortex, lateral prefrontal and frontopolar cortices, right temporoparietal regions, and the striatum. Notes: *: *P+*≥0.95 or ≤0.05; **: *P+*≥0.99 or ≤0.01; ***: *P+*≥0.999 or ≤0.001; *NS*: *P+*<0.95 or >0.05. ROI labels from the HCP multimodal parcellation (Glasser et al., 2016, *Nature*).

An interaction was also observed when predicting the immediate expected value (IEV; i.e., value of exploiting a familiar option) parameter. This was characterized by CTRL participants with increased behavioral apathy exhibiting heightened weighting of the IEV parameter (AMI Behave: *ρ=*0.46, *95%-CI*=0.03 to 0.80, *P+*=0.97; **Figure 4B**), which was stronger than the corresponding association across the TBI sample (*posterior difference, apathy-IEV correlation in CTRL versus TBI: behavioral AMI, P+=0.93*). Notably—greater IEV weighting on the Bandit task was associated with a reduced willingness to Accept high effort options in the Apples task across subjects (**Supplementary Figure 3**). This IEV by AMI behavior correlation in the CTRL group may therefore reflect a similar generalized bias towards low effort choices and greater apathy across tasks.

Behavioral and computational analyses suggest that apathetic TBI is characterized by a reduced drive to explore novel options. Model-based analysis of the fMRI data provided evidence about the underlying neural mechanisms. Specifically, this analysis centered on neural encoding of the RPE at feedback time. A wholebrain map found a distributed set of prefrontal, temporoparietal, cingulate, amygdala, and striatal regions that positively encoded RPEs, as well as negative RPE encoding in a region of dorsal anterior cingulate cortex (**Figure 4C**). We also found an interaction between emotional AMI scores and TBI group. Specifically, higher emotional AMI scores in TBI patients were associated with reduced RPE encoding in regions of the frontopolar cortex (FPC), dorsolateral prefrontal cortex (dlPFC), salience network (anteroventral and middle insula, rostral anterior cingulate), ventromedial prefrontal cortex (vmPFC), nucleus accumbens (NAC), and caudate (CAU; **Figure 4D**). In each case, they were driven by blunted RPE encoding as a function of apathy in TBI and a lack of correlation in CTRL (**Figure 4D**). These clusters scaled in the same direction with behavioral apathy, but only emotional apathy survived FLAME1 correction.

To formally link whole-brain RPE effects to computational parameters at the region-of-interest (ROI) level, we extracted post-hoc parameter estimates from a priori anatomical parcels from the Human Connectome Project (HCP) multimodal parcellation (i.e., the Glasser atlas^50^) corresponding to subgenual anterior cingulate cortex (sgACC; s32) and ventromedial FPC (area 10v). These regions were selected given their established role in driving apathy in TBI^2,44^, and their presence in the full group-level RPE encoding map (**Figure 4C**). In TBI patients, RPE encoding in s32 was positively associated with weighting the relative future value of exploration (BONUS; *ρ=*0.40, *95%-CI*=0.049 to 0.70, *P+*=0.978), but was unrelated to weighting of the immediate value of exploitation (IEV; *ρ*=0.18, *95%-CI*=−0.18 to 0.54, *P+*=0.831; **Supplementary Figure 4A-B**). s32 RPE encoding was also negatively associated with apathy, demonstrating a slightly stronger negative association with emotional relative to behavioral apathy (emotional: *ρ*=−0.38, *95%-CI*=−0.67 to −0.04, *P+*=0.021; behavioral: *ρ*=−0.32, *95%-CI*=−0.65 to 0.04, *P+*=0.049; **Figure 4D**). When partialling out AMI emotional apathy scores, the association between s32 RPE encoding and BONUS was no longer statistically credible (*ρ*=0.20, *95%-CI*=−0.20 to 0.56, *P+*=0.827).

A highly similar pattern of results was observed across s32 and frontopolar area 10v. There was a positive association between RPE encoding and BONUS weighting in area 10v (*ρ*=0.38, *95%-CI*=0.012 to 0.69, *P+*=0.974), but RPE encoding in 10v was not associated with IEV (*ρ*=0.15, *95%-CI*=−0.25 to 0.49, *P+*=0.773). 10v was also more strongly associated with behavioral rather than emotional apathy (*ρ*=−0.37, *95%-CI*=−0.67 to −0.012, *P+*=0.028; emotional apathy: *ρ*=−0.23, *95%-CI*=−0.59 to 0.12, *P+*=0.112). When partialling out AMI behavioral apathy scores, the strength of the positive association between 10v RPE encoding and BONUS was attenuated from strong to moderate evidence (*ρ*=0.31, *95%-CI*=−0.078 to 0.64, *P+*=0.934). Together, these data suggest that in the TBI group, reduced RPE encoding in sgACC and FPC during motivated decision-making is associated with both decreased directed exploration and increased apathy symptoms. Notably, partial correlation analyses revealed that the associations between s32 and 10v RPE encoding and directed exploration (i.e., BONUS) were tempered when controlling for concurrent levels of apathy—suggesting that individual differences in apathy contribute to their bivariate relationship.

### Post-hoc control analyses

We wanted to confirm that the observed associations between emotional apathy and diminished directed exploration in TBI were not confounded by depression or age-related change. First, we checked the presence or absence of associations between apathy (AMI subscales) and depression (BDI total scores). While behavioral apathy was strongly associated with depression (all subjects: *ρ*=0.62, *95%-CI* =0.42 to 0.78, *P+*=1.0; TBI only: *ρ*=0.71, *95%-CI*=0.49 to 0.87, *P+*=1.0), emotional apathy was not (all subjects: *ρ*=0.037, *95%-CI*=−0.26 to 0.34, *P+*=0.592; TBI only: *ρ*=0.071, *95%-CI*=−0.32 to 0.43, *P+*=0.636). There was also evidence that apathy and depression in TBI were more strongly associated in the behavioral relative to the emotional domain (*rho difference*=0.624, *95%-CI*=0.27 to 0.99, *P+*=0.998). This aligns well with prior findings that the emotional dimension of apathy is distinct from depression^57^.

Next, to confirm that our primary group-specific effects on directed exploration were not confounded by group differences in age or depression, we re-fit our primary Bayesian multi-level models including main effects of age and BDI, as well as an age*apathy interaction term. Importantly, the main effect of age (*b*=−0.072, *95%-CI*=−0.53 to 0.41, *P+*=0.370), the age*apathy interaction (emotional: *b*=0.020, *95%-CI*=−0.38 to 0.40, *P+*=0.542; behavioral: *b*=0.019, *95%-CI*=−0.20 to 0.27, *P+*=0.567), and the main effect of BDI (*b*=−0.045, *95%-CI*=−0.23 to 0.13, *P+*=0.305) were not credibly associated with BONUS weighting on the bandit task. The emotional apathy by TBI status interaction on BONUS was robust to the inclusion of age and BDI in the model (*b*=−0.764, *95%-CI*=-1.42 to −0.04, *P+*=0.019), whereas the behavioral apathy by TBI interaction was not (*b*=−0.261, *95%-CI*=−0.74 to 0.20, *P+*=0.138), which is unsurprising given the collinearity between behavioral apathy and depression.

Lastly, because the normative POMDP model demonstrated a moderately stronger fit to observed choice behavior in CTRL relative to TBI participants (*r*=0.79 versus *r*=0.69, *P+*=0.94), we determined whether overall model fit varied as a function of the critical apathy by group interactions. Emotional apathy was not credibly associated with POMDP model fit across TBI or CTRL participants (main effect: *b*=−0.088, *95%-CI*=−0.32 to 0.14, *P+*=0.213; AMI*group interaction term: *b*=−0.025, *95%-CI*=−0.32 to 0.27, *P+=*0.435). There was, however, a moderate interaction effect of behavioral apathy and group on POMDP fit (*b*=−0.131, *95%-CI*=−0.32 to 0.06, *P+=*0.083), which was driven by a positive trend in CTRL participants (*b*=0.089, *95%-CI*=−0.025 to 0.21, *P+=*0.938; consistent with heightened IEV exploitation in apathetic CTRL participants), versus a nonsignificant reduction in model fit associated with higher behavioral apathy in TBI (*b*=−0.044, *95%-CI*=−0.18 to 0.10, *P+=*0.259). Therefore, the association between emotional apathy and reduced directed exploration in TBI is independent of general TBI-related differences in the normativity of observed choices on the bandit task.

## Discussion

Does apathy reflect a reduced willingness to exert effort for known rewards, or a diminished motivation to explore new information for unknown *future* benefit? The present study dissociates these mechanisms, revealing that—while there is a general association between apathy and effort minimization across TBI and CTRL groups on both fMRI tasks—clinical apathy in TBI is uniquely characterized by decreased directed exploration to gain new information. These findings point to a dual-mechanism account of apathy, with specific implications for understanding and treating the motivational deficits that frequently follow TBI and other neurological disorders.

A failure to normatively weigh physical effort costs against the anticipated benefits of our actions has previously been linked to apathy across Parkinson’s Disease (PD), small vessel cerebrovascular disease^51^, and healthy control participants^13,14^. The current findings replicate this general pattern and extend it to the context of TBI-related apathy. Across both individuals with TBI and healthy controls, higher total apathy scores were related to a greater effort-based discounting of rewards. This behavior was mirrored by activity in a specific brain network involving somatomotor areas, midcingulate, middle insula, and the putamen. Crucially, cortico-insular and striatal circuits are known to encode subjective value expectations and the violation of those expectations during effortful tasks^52^. Our findings suggest that in more apathetic individuals, this network shows an exaggerated BOLD response to increasing effort demands. This hypersensitivity to effort costs may overwhelm the representation of potential rewards, providing a candidate neural mechanism for the observed increase in effort-based discounting in individuals with higher total apathy across both TBI and CTRL.

A core component of adaptive behavior in primates is curiosity—the drive to gain information, reduce uncertainty about the future, and seek novel experiences^53,54^. Our findings suggest that TBI-related apathy involves a specific disruption of this process. Behaviorally, while both TBI and CTRL participants learned to exploit familiar rewarding options, individuals with TBI and higher apathy showed a reduced motivation to engage in directed exploration of novel, uncertain choices. Our model-based fMRI analysis identified a clear neural correlate for this deficit: a blunted reward prediction error (RPE) signal at the time of feedback. This RPE signal was attenuated in apathetic TBI patients across a network including the frontopolar cortex (FPC), ventromedial prefrontal cortex (vmPFC), and striatum. vmPFC and striatum encoding of RPEs aligns well with recent meta-analyses of RPE encoding in humans^55,56^. These effects were in the same direction for both behavioral and emotional apathy—but were particularly robust with respect to emotional apathy. Recent machine learning-based deconstruction of the apathy phenotype found that emotional apathy is unique relative to the other domains, and is the only AMI domain that *is not* associated with depression or anhedonia^57^. This dissociation was replicated in the current study, where depression correlated strongly with behavioral but not emotional apathy in TBI and across the full sample. This study also found that emotional apathy was associated with a core deficit in processing the *intensity* of emotionally salient information^57^. This also accords with current results—diminished RPE encoding as a function of increased emotional apathy suggests that individuals with this symptom have a decreased neural encoding of the relative *intensity* of rewarding feedback. From a translational perspective, this finding suggests that emotional amotivation may be a particularly informative domain for screening underlying decision-making deficits post-TBI, and future precision-medicine studies should evaluate whether using emotional apathy scores is beneficial for stratifying patients for interventions targeting dopaminergic RPE circuits.

The finding that this signal is blunted in FPC and subgenual cingulate (areas 10v and s32, respectively) in apathetic individuals with a diminished motivation to engage in directed exploration—and that RPE encoding in both regions positively scales with exploration BONUS in TBI—is particularly noteworthy. A ventromedial prefrontal cortex cluster covering both ventral FPC and s32 was previously linked to apathy in penetrating TBI patients via voxel-based lesion-symptom mapping (VLSM)^2^, and resting-state functional connectivity between the mask derived from that VLSM and dorsal anterior cingulate cortex was found to scale with apathy in patients with closed-head TBI^44^. Whereas subgenual cingulate dysfunction is causally linked to mood dysregulation in idiopathic depression^58–60^, structural and functional changes in this visceromotor valuation region post-TBI may instead blunt the affective feedback signals needed to motivate goal-directed behavior^cf.61^, contributing to emotional apathy.

The FPC is not as commonly linked to affective valuation as sgACC, but existing data from healthy volunteers provides clear evidence for its role in the cognitive architecture of goal-directed exploration. Not only is FPC critical for exploration and other forms of information-seeking^25,62–65^, but it is also involved in the ‘supervised control’ required to monitor progress through a sequence of tasks that are not habitual or low effort^66,67^. Blunted RPE encoding in the FPC may therefore reflect a dual deficit: it is associated with an impaired ability to learn the value of exploration, alongside a disrupted capacity to monitor when a strategic shift away from simple exploitation is necessary, leading to the inertia characteristic of apathy. This is consistent with recent meta-reinforcement learning (meta RL) models, which propose that RPEs support the updating of behavioral policies that guide adaptive information seeking and uncertainty reduction^68^. Altogether, we interpret the current findings as preliminary support for a latent cascade—whereby TBI-related RPE blunting in s32 and 10v impairs directed exploration (BONUS), which in turn manifests clinically as apathy, explaining why partialling out apathy symptoms attenuates the direct brain–behavior link. That said, larger longitudinal samples are required to formally test this mediation pathway. This circuit disruption could potentially play a role in driving comorbid motivated choice pathologies commonly observed post-TBI—including TBI-associated increases in addiction^69^ and obsessive-compulsive disorder^70^.

At first, the current findings may appear to contrast with recent work by Gilmour and colleagues (2024) on explore-exploit decision-making and apathy in Parkinson’s Disease (PD)^16^. Using a bandit task, Gilmour et al. observed reduced value-based learning as a function of apathy in PD, driving apathetic PD patients to make more noisy decisions, otherwise known as ‘random exploration’. Notably, Gilmour et al. used a four-armed ‘Restless Bandit Task’—where reward probabilities were continuously changing, making learning to exploit familiar rewards a cognitively demanding task. Our ‘Novelty-Bandit Task,’ in contrast, was designed so that exploiting the best familiar option was a stable, low-effort strategy. In fact, increased weighting of the value of exploitation in our task was associated with effort discounting, and increased in individuals with subclinical apathy. The more cognitively effortful component of our Novelty-Bandit paradigm is the tracking of uncertainty in the reward environment, to evaluate the potential informational gain of exploration (i.e., ‘directed exploration’). Hence, while Gilmour et al. found apathy in PD is associated with more random exploration, our findings point to a distinct computational profile of diminished exploration in apathetic TBI patients. Specifically, our results indicate that—in a choice task where learning to exploit requires *less* cognitive effort than the difficult decision to explore novel options^cf., 71^—clinically-acquired apathy may be more related to a diminished ability to track when it would be beneficial to explore new information for unknown future benefit (i.e., less effective directed exploration). The apparent discrepancy between our results and those of Gilmour et al. (2024) may therefore reflect differences in task demands—while restless bandits probe cognitively effortful learning to exploit under shifting reward contingencies, in our task, learning to exploit is relatively easy, making it better suited to isolating the goal-directed drive to explore novel options for latent future benefit.

Consistent with previous reports^4,72,73^, our TBI cohort exhibited elevated levels of chronic post-concussive symptoms, depression, anxiety, and impulsivity compared to controls. Furthermore, within the TBI group, repeated injuries exacerbated neurobehavioral symptoms, anxiety, and impulsivity, aligning with evidence suggesting cumulative effects can be more impactful than single injury severities for certain chronic psychiatric outcomes^1^. Crucially, however, apathy severity itself, measured using both the AES and AMI scales, was elevated across the TBI spectrum but did not significantly vary with injury repetition or severity (mTBI vs. msTBI) in our sample. This underscores apathy as a prevalent and potentially distinct consequence of TBI, necessitating dedicated mechanistic investigation beyond simply indexing injury characteristics.

Some limitations of the current study must be considered. First, the lack of longitudinal assessments and absence of any functional outcome measurements limited our ability to directly link our laboratory-based findings to real-world symptom trajectories and clinical outcomes. More work is needed to fully realize the potential translational impact of these findings. In addition, the use of a hypothetical effort-value tradeoff in the scanner—rather than use of a dynamometer to record fMRI responses during extended isometric muscle contraction in the scanner—means that the Apples task was more sensitive to anticipated, rather than actual, willingness to exert physical effort. While this is a common and well-validated approach, future studies using fMRI paradigms that enable the measurement of neural responses to experienced effort and TBI-associated apathy are warranted. Lastly, while the current sample size represents a reasonable recruitment effort for a clinical neuroimaging study with multiple high trial count model-based fMRI tasks, future larger-scale studies are needed to further establish the sensitivity and specificity of blunted RPE for predicting reduced exploration and increased emotional apathy in TBI.

This study provides novel evidence dissociating two distinct neurocomputational mechanisms contributing to apathy following TBI. A specific deficit in computing RPEs during motivated choice was closely associated with the inability of individuals with acquired apathy following a TBI to flexibly explore novel and potentially beneficial choice options. These findings have significant implications for understanding and potentially treating apathy after TBI. They move beyond characterizing apathy simply as reduced willingness-to-exert effort or blunted reward valuation, providing a distinct computational target: The prospective devaluation of uncertain choice options that may be beneficial in the future. Future behavioral, pharmacologic, and neurostimulation based interventions for apathy should therefore consider focusing on normalizing the flexible exploration of novel and unfamiliar choice options in patients with TBI.

## Data Availability

All data produced in the present study are available upon reasonable request to the authors.

## Acknowledgements

While conducting this work, JH was supported by National Institutes of Health through the National Institute of General Medical Sciences award number P20GM109089, the National Institute on Alcohol Abuse and Alcoholism award number R01AA030283, and the National Science Foundation award number #2237795. We would like to thank the UNM Center for Advanced Research Computing, supported in part by the National Science Foundation, for providing the research computing resources used in this work.

## Disclosures

All authors report no biomedical financial interests or potential conflicts of interest.

## Footnotes

1 Note: *N*=2 TBI participants met more than one exclusion criterion (i.e., there are 5 additional excluded subjects, despite 7 total reasons for exclusion listed).

## Notes

### Competing Interest Statement

The authors have declared no competing interest.

### Author Declarations

University of New Mexico Health Sciences Center, Human Research Protections Office, #19-053

### Summary of Updates

Results and discussion were updated significantly thanks to some constructive feedback from two anonymous reviewers.

