## Supplementary figures and images for "Effort aversion and diminished exploration in apathy associated with Traumatic Brain Injury"

### Supplemental Figure 1

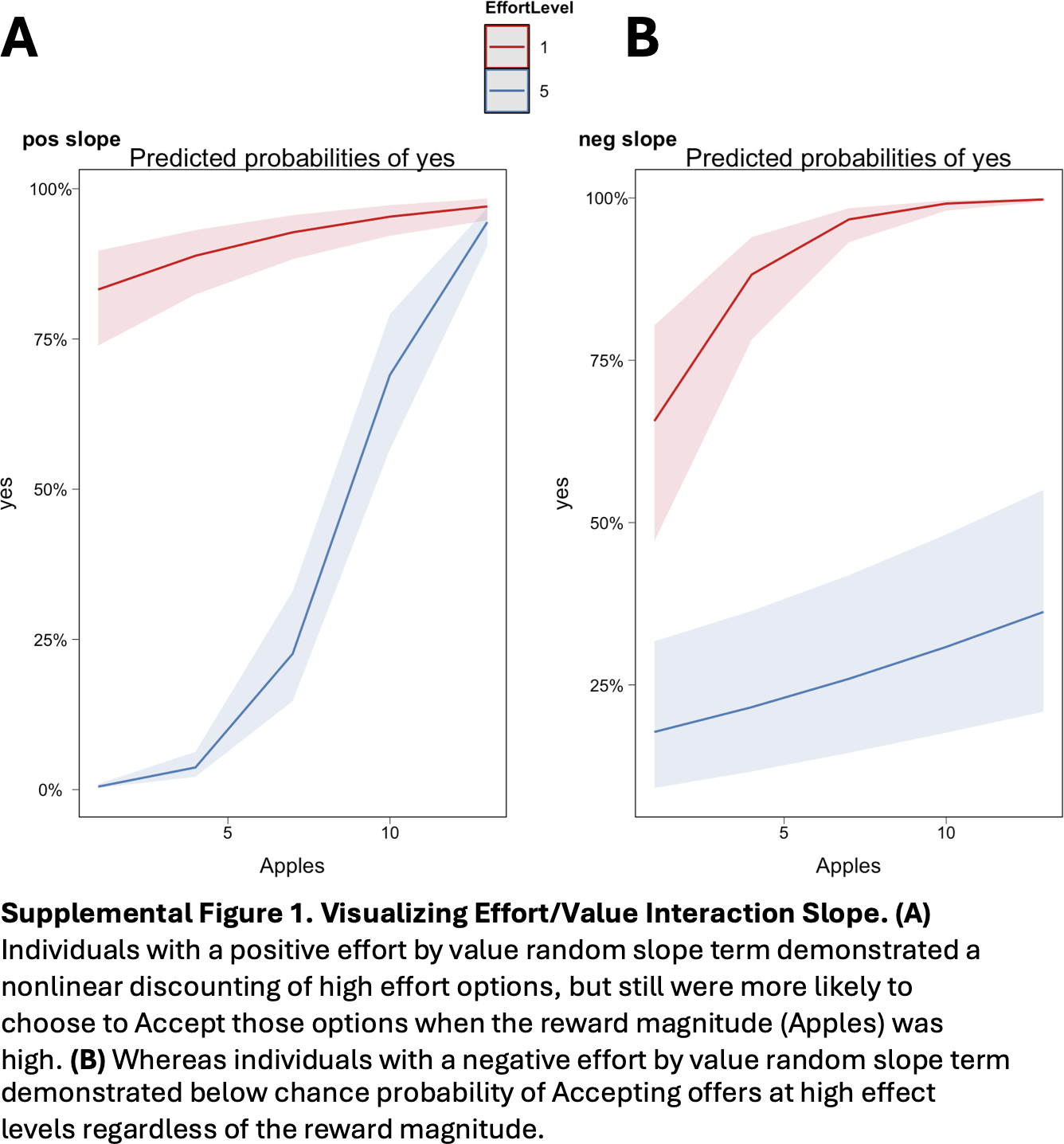

### Supplemental Figure 2

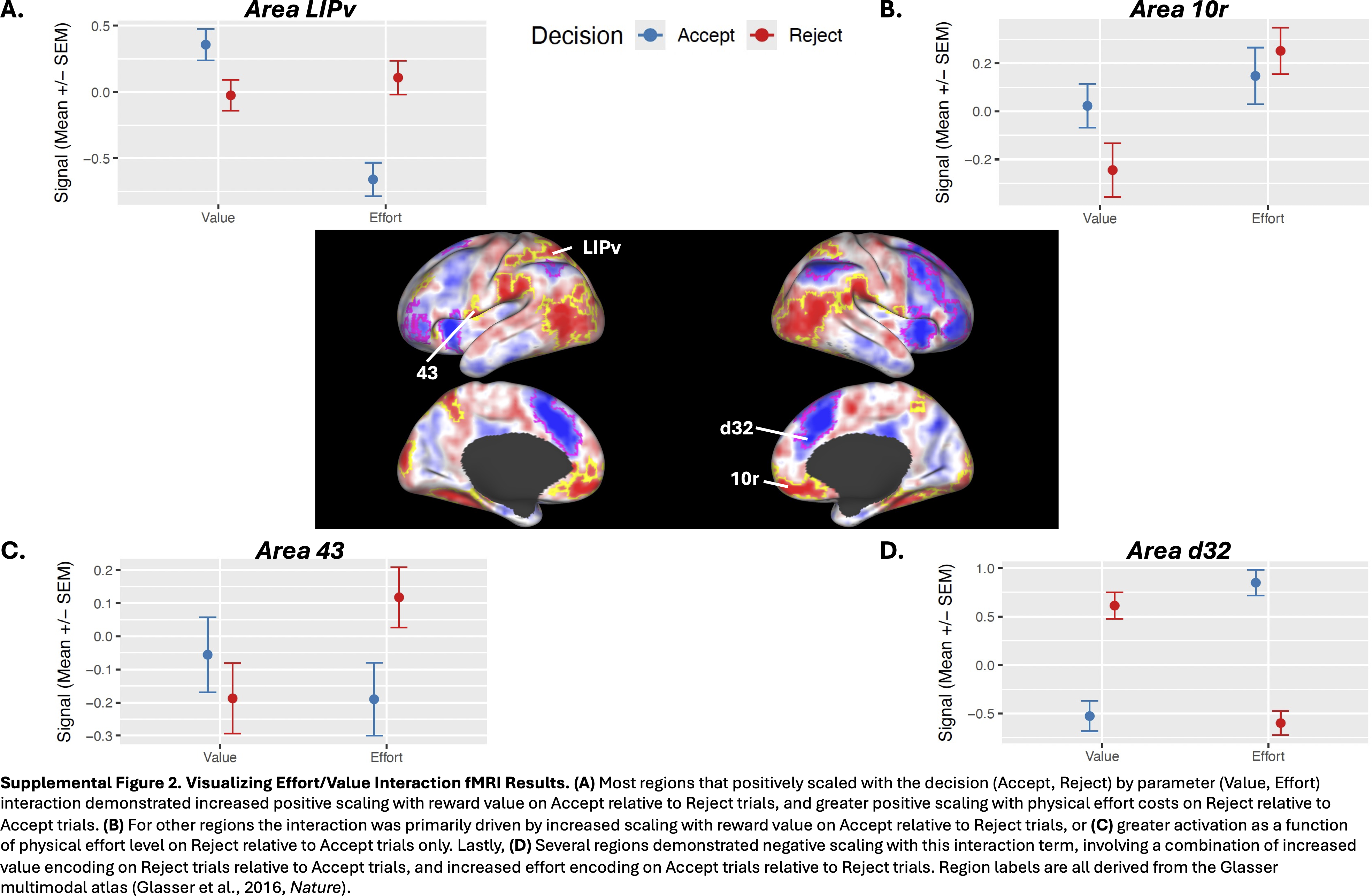

### Supplemental Figure 3

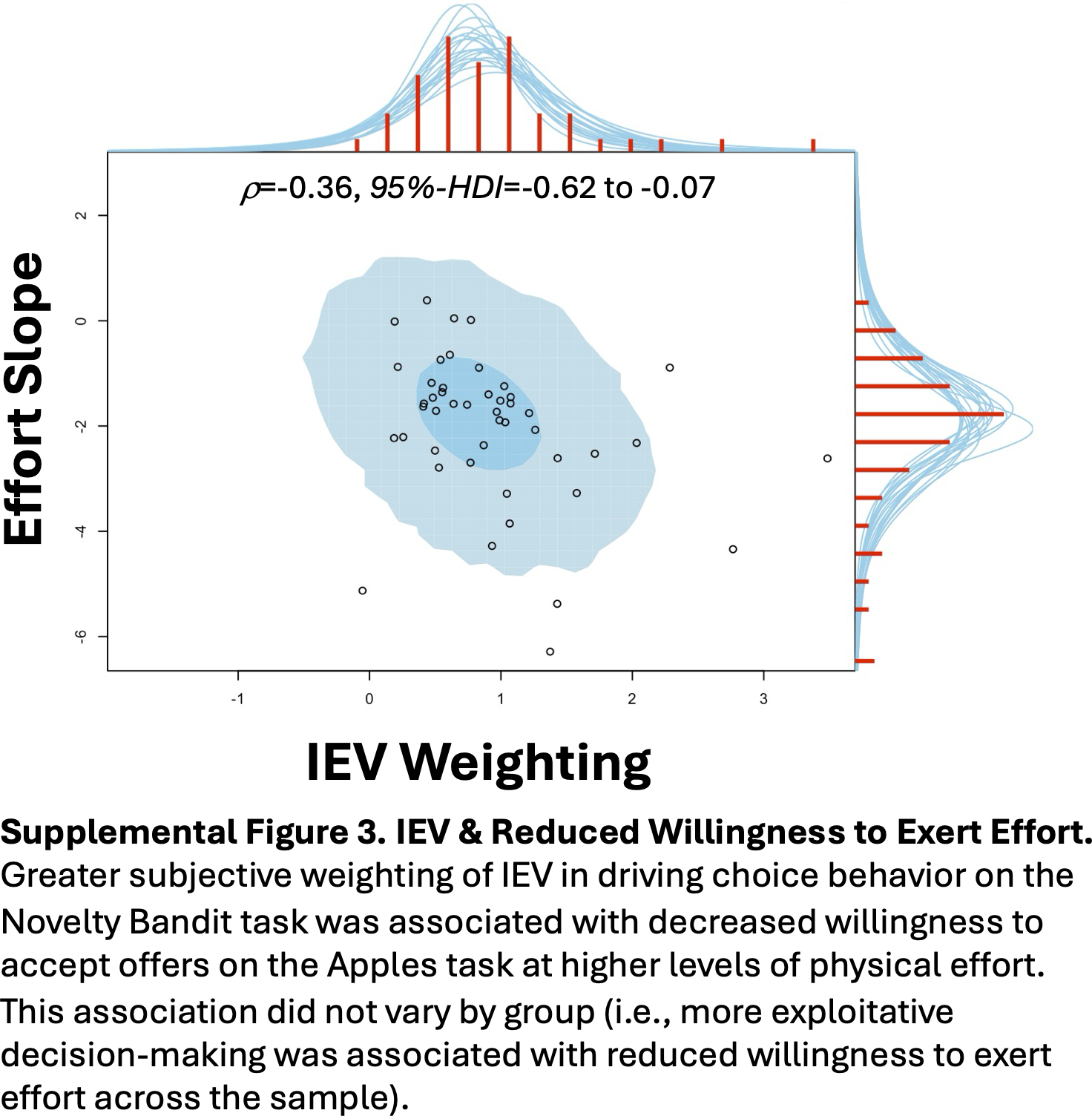

### Supplemental Figure 4

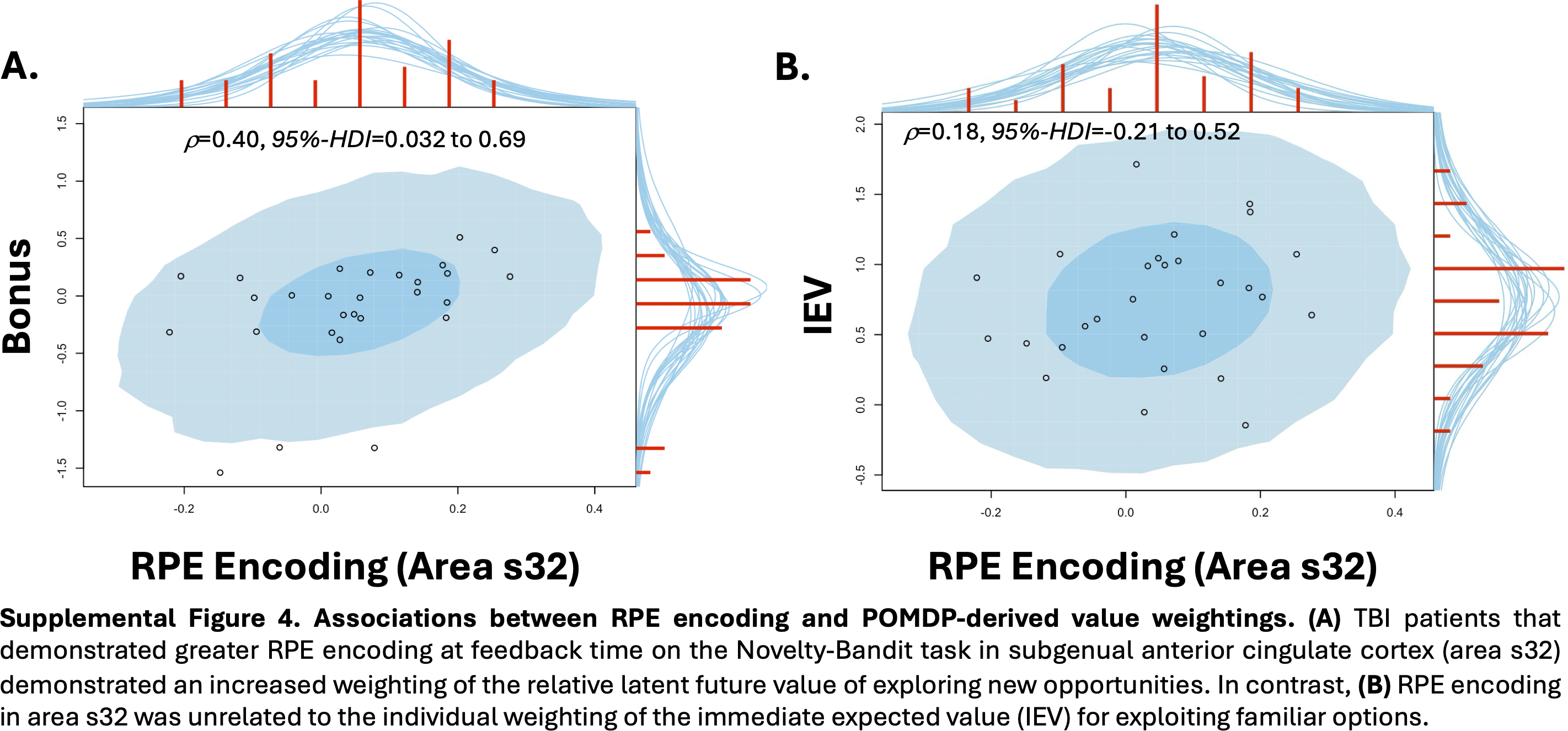
